# Smart Pooled sample Testing for COVID-19: A Possible Solution for Sparsity of Test Kits

**DOI:** 10.1101/2020.04.21.20044594

**Authors:** Syed Usama Khalid Bukhari, Syed Safwan Khalid, Asmara Syed, Syed Sajid Hussain Shah

## Abstract

There is an exponential growth of COVID-19. The adaptation of preventive measures to limit the spread of infection among the people is the best solution to this health issue. The identification of infected cases and their isolation from healthy people is one of the most important preventive measures. In this regard, screening of the samples from a large number of people is needed which requires a lot of reagent kits for the detection of SARS-CoV-2. The use of smart pooled sample testing with the help of algorithms may be a quite useful strategy in the current prevailing scenario of the COVID-19 pandemic. With the help of this strategy, the optimum number of samples to be pooled for a single test may be determined based on the total positivity rate of the particular community.

## Introduction

Over the past few months, COVID-19 has attacked more than two million people around the globe and it has caused more than one hundred and fifty thousand deaths. The rapid spread of COVID-19 occurred due to rapid transmission from infected to healthy persons. The causative agent of Coronavirus disease (COVID-19) has been named as severe acute respiratory syndrome coronavirus 2 (SARS-CoV-2). Person to person transmission of SARS-CoV-2 as this pathogen is present in the respiratory droplets of the infected person and respiratory droplets containing viruses may spread the virus to healthy persons [1-2]. Some persons infected with severe acute respiratory syndrome coronavirus 2 (SARS-CoV-2) may have a mild disease or may be asymptomatic [3-4]. The apparently healthy people with COVID-19 can be a source of the spread of SARS-CoV-2 to the community [5]. The detection of such cases in the community has got vital importance in the prevention of viral spread. The mass level screening of the community for the identification of infected cases is a very important step to control the damage to the health of the community.

For the detection of COVID-19, the PCR test is performed on the specimens acquired from the nasal swab of asymptomatic person and bronchial secretion of severely ill patients. This test is not routinely performed in all laboratories and it required special instruments and specific expertise along with specific kits for the detection of severe acute respiratory syndrome coronavirus 2 (SARS-CoV-2) in patients of COVID-19.

There is less availability of kits and diagnostic facilities for the vast majority of the people around the world. The smart utilization of resources may be of vital importance in controlling the pandemic. In this regard, the possibility of pool sample testing may be explored which may be helpful in reducing the consumption of less available kits of this pathogen and it may also reduce the workload over the overburdened laboratory staff.

Data on COVID-19 testing from various countries revealed that in the mass testing for the screening purpose, the majority of the test is negative. The pooling of the negative sample will reduce kit consumption without affecting the outcome. The possibility of mixing the positive specimen with the negative may result and it would affect the true results. The one possibility is that the positive result may become negative due to the dilution with the negative specimen. The PCR can detect the small number of pathogens and the specimen from positive patient usually contains a large number of viral particles, so the possibility of the positive specimen is to become negative by mixing with the negative specimen is very remote. The studies revealed that there is no significant effect of the pooling of negative specimens with the positive specimen in terms of the detection of the positive test by PCR methods [6]. Another study showed that the positive specimen may be detected even when it is diluted with up to 64 specimens [7].

The other limitation is that the mixing of one positive with a negative specimen will reveal positive results, and for the identification of the true positive patient, the tests will have to be repeated on all the specimens and it may consume more kits. To address these issues, the application of algorithms after gathering the data of previous test results from that country may be quite helpful in reducing the possible consumption of test kits.

The aim and objective of the present article are to find out the number of samples to be pooled in a particular community for the reduction of utilization of kits.

## Materials & Methods

In this study, we work on a research question that if we group people and perform the test for COVID-19 on a group, can we achieve any optimization in the utilization of recourse (in our case are test kits) required to perform tests.

To answer that question, we develop a hypothesis that if we made a group with the number of people that is some power of 2, and apply divide and conquer strategy, a significant optimization can be achieved. Following are the steps to perform divide and conquer strategy.

### Steps

1. Get the group size ***n***
2. Perform a test on the whole group.
3. If the result of the whole group is negative, then infer that everyone in the group has a negative result.
4. If the result of the group is positive, it means that one or more than one member of the group has a positive result.
5. Divide the group into two equal subgroups. As ***n*** should be of the power of ***2***, hence dividing the group into two equal subgroups is possible.
6. Go to step 2 for both groups in parallel.

*Let take an example, suppose we made a group of 8 (2*^*3*^)

**Figure 1:**
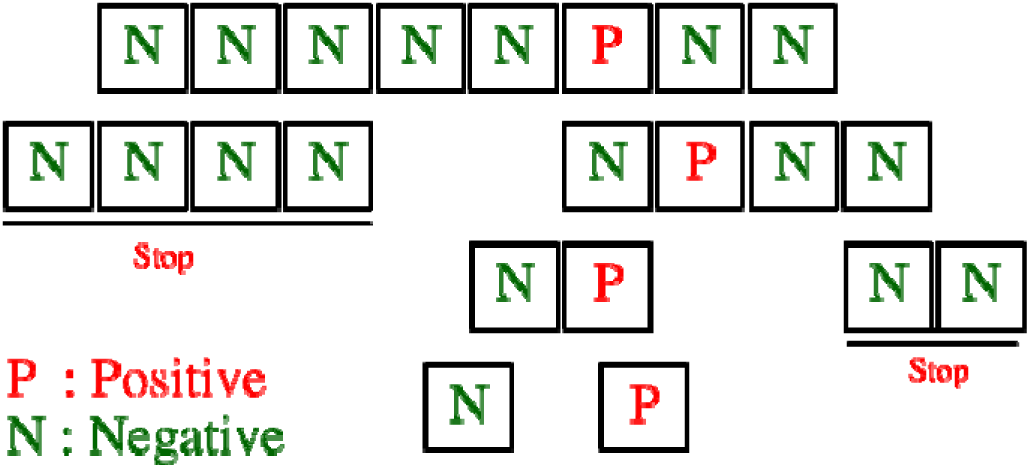
visual representation of divide and conquer strategy.

As the tests for COVID-19 are very critical and we need 100% surety that no one infected by it, can skip because of basic assumptions of divide and conquer strategy. Hence the standard calculation of divide and conquers can’t be used. Therefore, we derived the formulae to give us approximate maximum tests needed to perform to be 100% sure everyone is tested in best, average and worse case scenarios.

*Let* ***n*** *is the total number of people in one group*.

### Best case scenario

The best-case scenario is the scenario in which no one in the group have COVID-19. Which means that the whole group’s result will come negative. In this scenario, we will need no further tests.

*In the best-case scenario, total tests need to be done are*

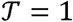

### Worst Case Scenario

In this scenario, 40 % or more people of the group are infected by SARS-CoV-2, and the samples of these people are uniformly distributed in the group.

*For example, if* ***n*** *= 8*

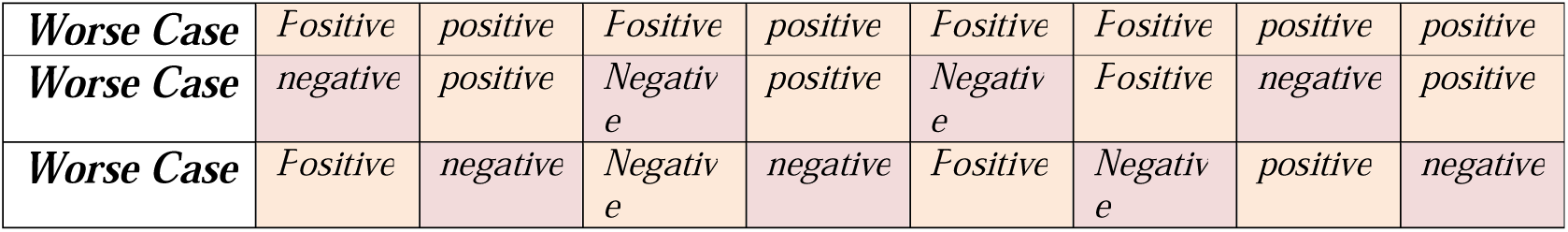

|  |  |  |  |  |  |  |  |  |
| --- | --- | --- | --- | --- | --- | --- | --- | --- |
| **Worse Case** | Positive | positive | Positive | positive | Positive | Positive | positive | positive |
| **Worse Case** | negative | positive | Negative | positive | Negative | Positive | negative | positive |
| **Worse Case** | Positive | negative | Negative | negative | Positive | Negative | positive | negative |

*In the worst-case scenario, total tests need to be done are*

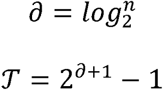

### Average Case Scenario

All the other possible scenarios, which do not fall in best or worse case scenarios, fall into average case scenarios.

*In the average-case scenario, total tests need to be done are*

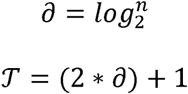

### Exceptions

For a group of 2, the following steps should be taken.

- Check the whole group.
- If the combined group result is positive then perform the test on only one sample.
- If his/her result is negative, then there is no need to perform the test on the other sample. Where if his/her result is positive, then it is mandatory to perform the test on the other sample as we are not sure if the other is positive or not.

## Results

As the scenarios are dependent upon the percentage of people who are infected by SARS-CoV-2. Hence the best way to reach a logical conclusion was to find the ratio between the number of tests conducted by any country, and how many people were reported positive. In this study, we are going to refer this term as Test to Positive Ration (TPR).

**Table 1.** Shows TPR of 6 countries (https://www.worldometers.info/coronavirus/)

| COUNTRY | TOTAL TEST | POSITIVE REPORTED | TPR |
| --- | --- | --- | --- |
| USA | 2558253 | 506008 | 19.8 |
| ITALY | 906864 | 147,577 | 16.3 |
| AUSTRALIA | 351380 | 6303 | 1.8 |
| TURKEY | 307210 | 47029 | 15.3 |
| UK | 334974 | 78991 | 23.6 |
| PAKISTAN | 57836 | 4970 | 8.6 |

For a better understanding, analyzing and to find the most optimal group set, we developed a simulator. The job of the simulator is to generate a random population with given TPR as input, and then using different group sizes perform the divide and conquer strategy, and give results.

### All the analysis is done using the concept that 1 test kit can do a test of one patient. The Simulator takes the following inputs

|  |  |
| --- | --- |
| **Population size** | We used 10,000 |
| **TPR** | we used the TPR of the countries given in table 1 |
| **Group size** | we used different group sizes 2 till 32 |

A group of 2 is a special case, therefore in the simulator, it is treated as a special case and calculation is done as per requirements.

we generated 100, different populations of 10,000 and then simulate them. The average results are shown in graph 1. The results are in percentage showing how much tests we can save to tests a population of 10,000.

**Graph 1:**
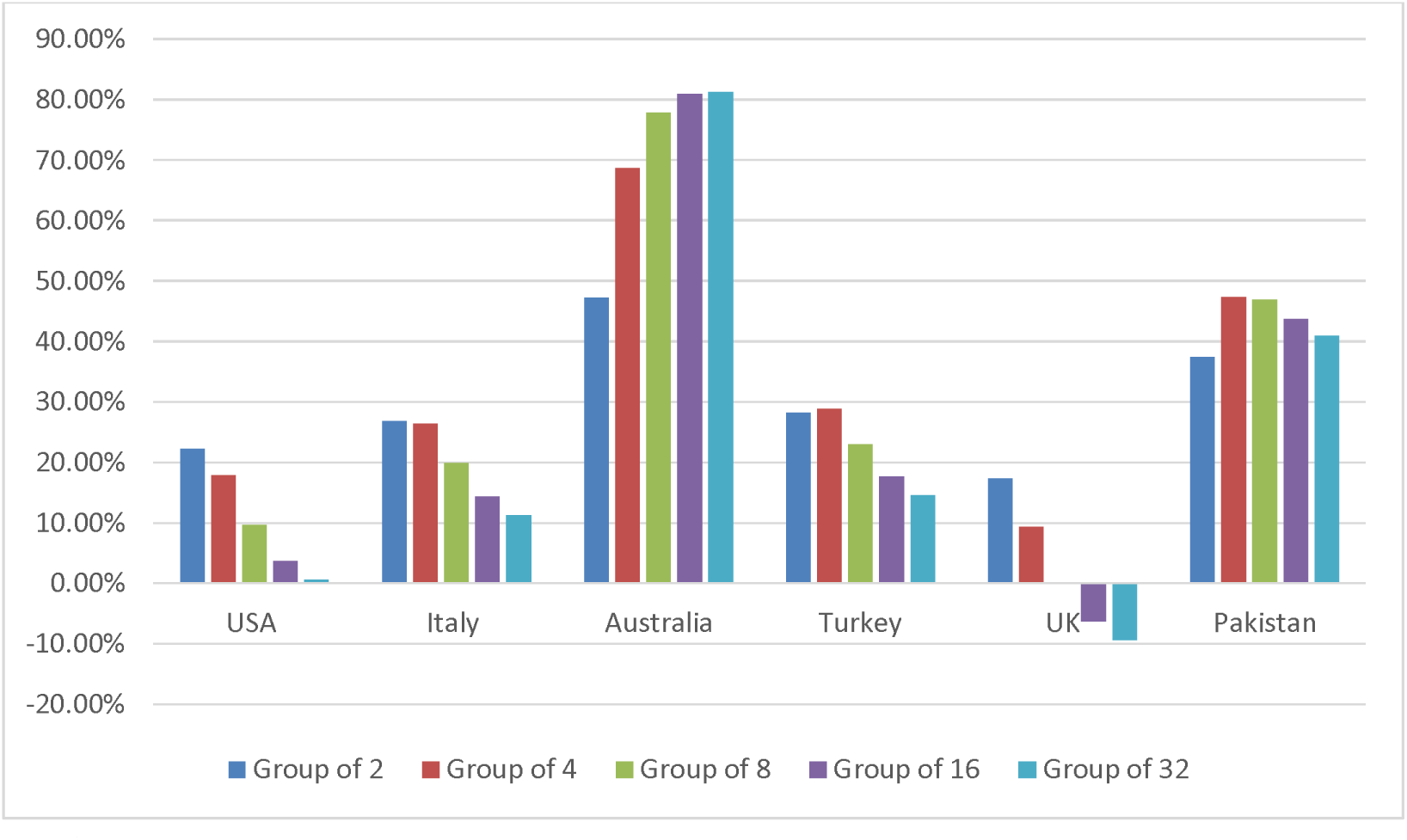
Showing how many tests in percentage, every country can save to tests a population of 10,000.

**Table 2:**
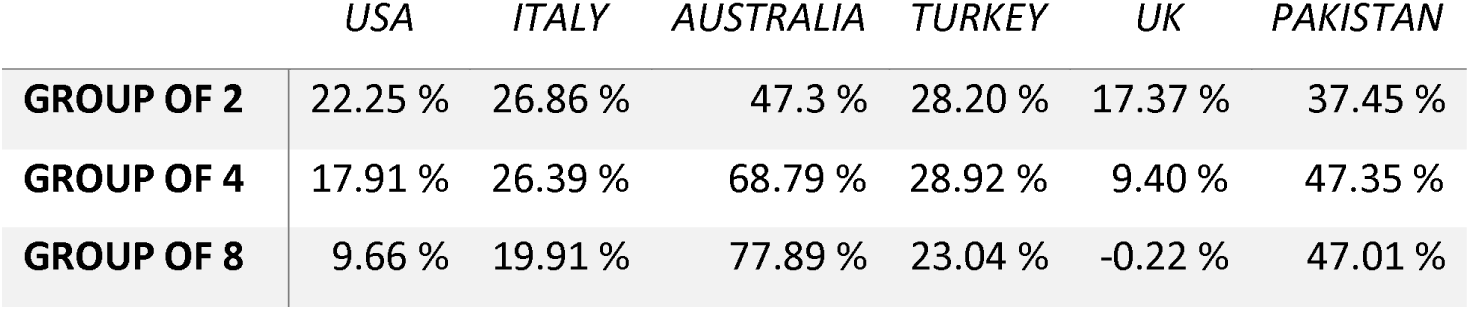
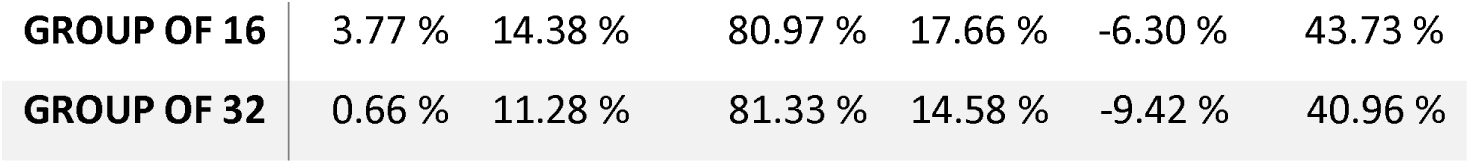
Results of simulation

|  | USA | ITALY | AUSTRALIA | TURKEY | UK | PAKISTAN |
| --- | --- | --- | --- | --- | --- | --- |
| **GROUP OF 2** | 22.25 % | 26.86 % | 47.3 % | 28.20 % | 17.37 % | 37.45 % |
| **GROUP OF 4** | 17.91 % | 26.39 % | 68.79 % | 28.92 % | 9.40 % | 47.35 % |
| **GROUP OF 8** | 9.66 % | 19.91 % | 77.89 % | 23.04 % | -0.22 % | 47.01 % |
| **GROUP OF 16** | 3.77 % | 14.38 % | 80.97 % | 17.66 % | -6.30 % | 43.73 % |
| **GROUP OF 32** | 0.66 % | 11.28 % | 81.33 % | 14.58 % | -9.42 % | 40.96 % |

**Table 3:**
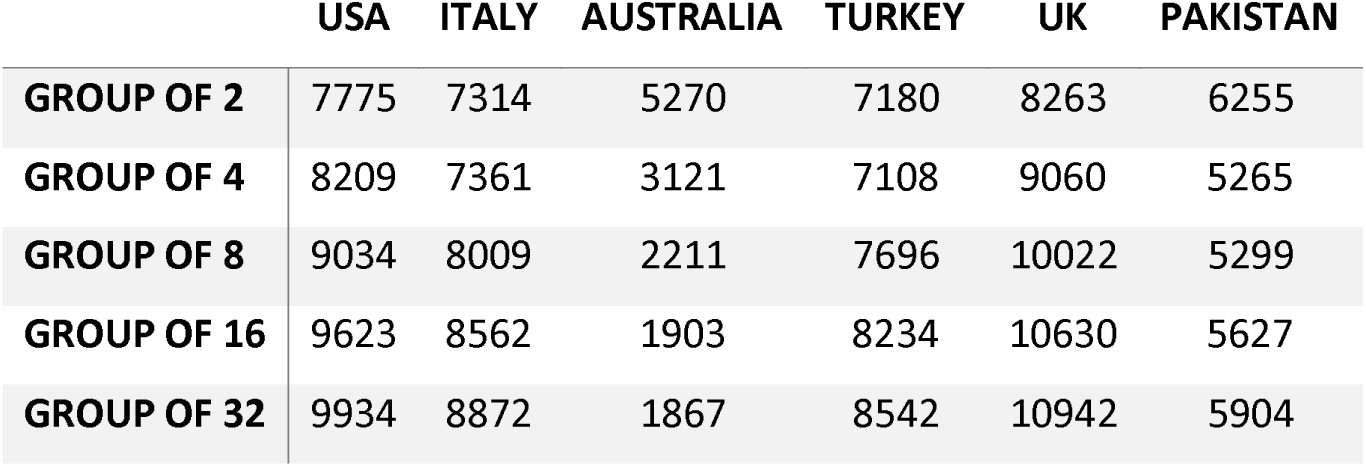
Minimum number of tests needed to test a population of 10,000

The simulation results show that if the TPR is directly linked with the number of tests needed to test a population of 10,000. It TPR is low, required tests will be low and if TPR is high then required tests will be high.

To get a better understanding of the TPR, the number of tests needed and which group size will be optimal for which TPR. We perform the number of tests on 100 different populations of size 10,000 using different TPRs.

**Graph 2:**
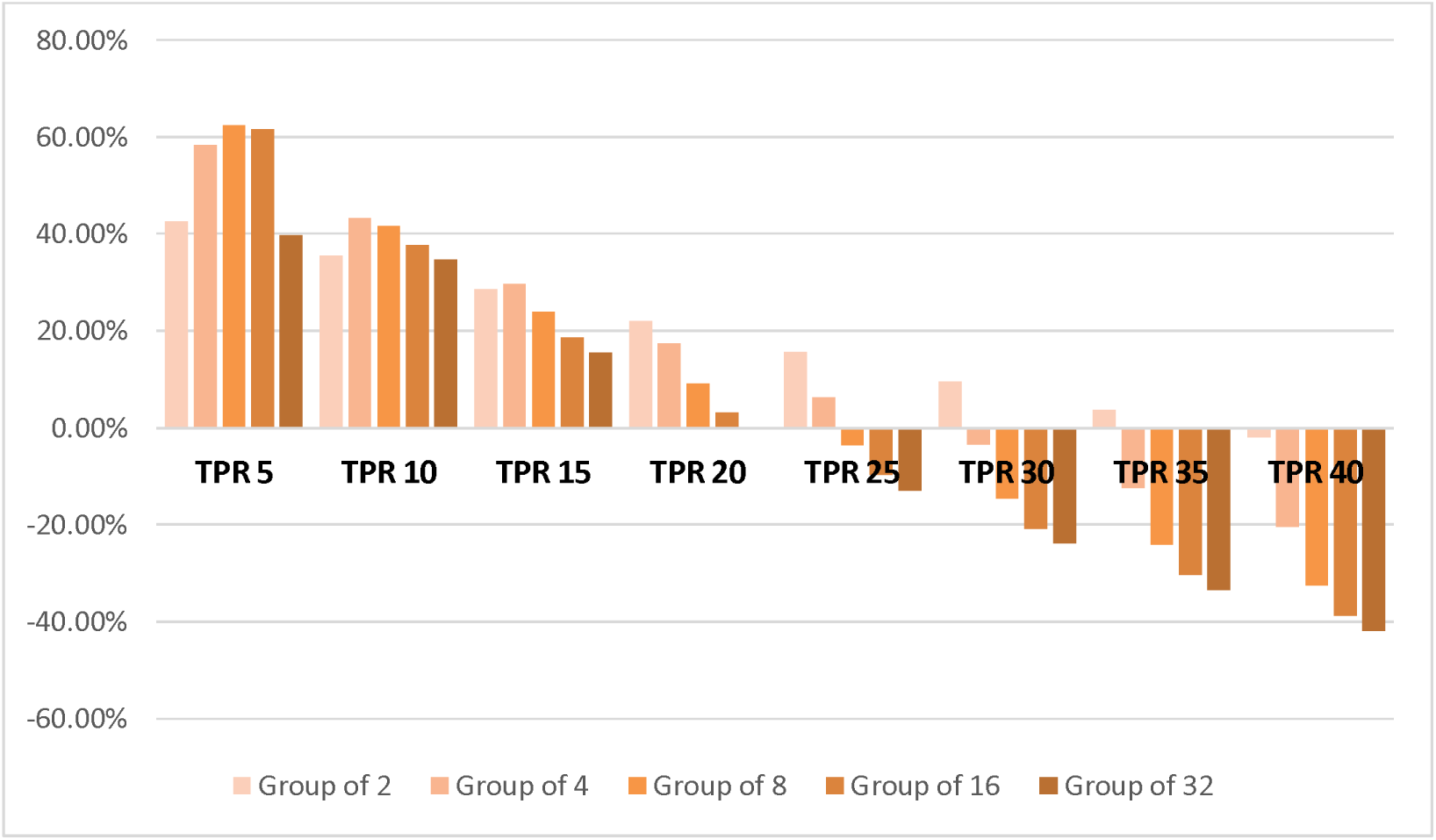
Showing how many tests in percentage, one can save to tests a population of 10,000.

As results are shown in graph 2 and table 4, one can conclude that if the TPR is below 30, a significant optimization can be achieved resulting in performing fewer tests for every 10,000 population. The results also show that if the TPR is below or close to 10, higher group size is more beneficial. Where group size of 2 might be a better choice if TPR is higher than 15. It is also concluded that if TPR is above 35 one should not use this grouping method as it will add extra burden and result in more tests.

**Table 4:**
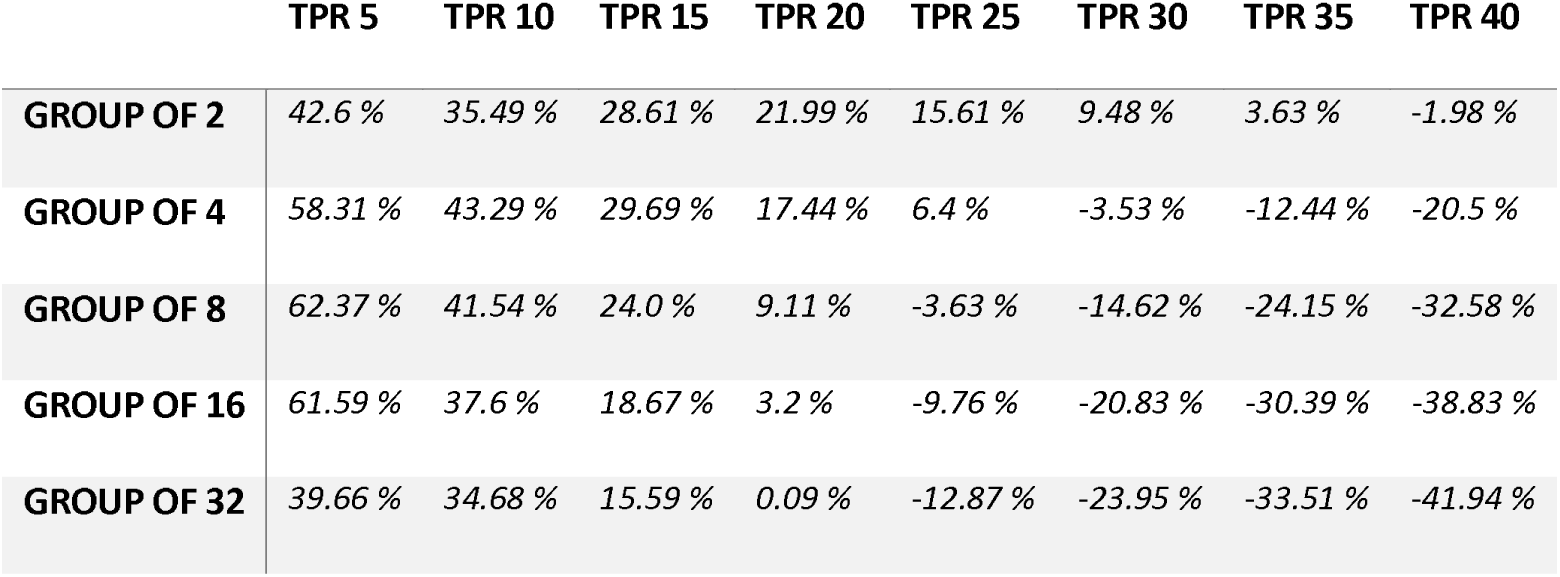
Number of test (in percentage), one can save to test a population of 10,000 with respect to TPR

|  | TPR 5 | TPR 10 | TPR 15 | TPR 20 | TPR 25 | TPR 30 | TPR 35 | TPR 40 |
| --- | --- | --- | --- | --- | --- | --- | --- | --- |
| **GROUP OF 2** | 42.6 % | 35.49 % | 28.61 % | 21.99 % | 15.61 % | 9.48 % | 3.63 % | -1.98 % |
| **GROUP OF 4** | 58.31 % | 43.29 % | 29.69 % | 17.44 % | 6.4 % | -3.53 % | -12.44 % | -20.5 % |
| **GROUP OF 8** | 62.37 % | 41.54 % | 24.0 % | 9.11 % | -3.63 % | -14.62 % | -24.15 % | -32.58 % |
| **GROUP OF 16** | 61.59 % | 37.6 % | 18.67 % | 3.2 % | -9.76 % | -20.83 % | -30.39 % | -38.83 % |
| **GROUP OF 32** | 39.66 % | 34.68 % | 15.59 % | 0.09 % | -12.87 % | -23.95 % | -33.51 % | -41.94 % |

## Discussion

COVID-19 has been declared a pandemic by WHO. Until now, there is no proven effective medicine against the COVID-19. The best solution is to adopt preventive measures to limit the spread of infection among the people. One of the important measures is to identify infected cases and isolate them from healthy people. In this regard, screening of the samples from a large number of people is required. For the diagnosis of SARS-CoV-2 infection, the PCR test is done on the specimens obtained from the upper respiratory tract (swabs from nasopharynx, oropharynx or aspirates from nasopharynx) or the lower respiratory tract (sputum, endotracheal aspirate, and bronchoalveolar lavage). The highest positive rate is seen in bronchoalveolar lavage specimens [8].

With the spread of COVID-19, There will be an increase in the requirement of kits for the detection of SARS-CoV-2. As the availability of test kits for SARS-CoV-2 are limited, the possibility of pool sample testing may be adapted to overcome the shortage of kit reagents. In the pool testing, the multiple specimens from different suspected cases are combined to perform them in a single test process, and this method is helpful in the improvement of a screening program for the community screening [9-10]. The important thing is pool sample testing is the number of tests to be included in the sample pool. If this number is not optimum, then there may be more utilization of test reagents and it would enhance the wastage of resources and efforts. The determination of an optimum number of specimens for the pooled sample testing is determined by the application of algorithms as discussed in the results. This application determines the optimum number of tests to be pooled for a single test and it will depend upon the total positive rate of COVID-19 cases in that particular community.

If the TPR is below 30, a significant optimization can be achieved resulting in performing fewer tests for every 10,0000 population. Results show that if the TPR is below or close to 10, higher group size is more beneficial. Similarly, a group size of 2 would be appropriate if TPR is 20 to 35, but in case TPR is more than 35 then the use of this grouping method would not be feasible, rather as it will add extra burden and cost.

## Conclusion

The application of algorithms to determine the appropriate number of specimens to be pooled for a single test would be a very cost-effective solution for the screening of community for the diagnosis of COVID-19.

## Data Availability

https://www.worldometers.info/coronavirus/

